# Comparing azithromycin to amoxicillin in the management of uncomplicated severe acute malnutrition in Burkina Faso: a pilot randomized trial

**DOI:** 10.1101/2021.06.15.21258967

**Authors:** Kieran S. O’Brien, Ali Sié, Clarisse Dah, Millogo Ourohire, Moussa Ouédraogo, Valentin Boudo, Ahmed M. Arzika, Elodie Lebas, Fanice Nyatigo, William W. Godwin, J. Daniel Kelly, Benjamin F. Arnold, Catherine E. Oldenburg

## Abstract

**Introduction:** Given the potential for asymptomatic infection in children with uncomplicated severe acute malnutrition (SAM), the World Health Organization recommends a broad-spectrum antibiotic like amoxicillin. Azithromycin is a promising alternative to amoxicillin as it can be administered as a single dose and has efficacy against several pathogens involved in the burden of infectious disease and mortality in this population. In this pilot study, we aimed to establish the feasibility of a larger randomized controlled trial and to provide preliminary evidence comparing the effect of azithromycin to amoxicillin on weight gain in children with uncomplicated SAM.

**Methods:** This pilot randomized trial enrolled children 6-59 months old with uncomplicated SAM at 6 healthcare centers in Burkina Faso. Participants were randomized to a single dose of azithromycin or a 7-day course of amoxicillin. All participants received ready-to-use therapeutic food and were followed weekly until nutritional recovery and again at 8 weeks. The primary feasibility outcomes included enrollment potential, refusals, and loss to follow-up. The primary clinical outcome was weight gain (g/kg/day) over the 8-week period. Outcome assessors were masked.

**Results:** Between June and October 2020, 312 children were screened, 301 were enrolled with 0 refusals, and 282 (93.6%) completed the 8-week visit. Average weight gain was 2.5 g/kg/day (SD 2.0) in the azithromycin group and 2.6 (SD) 1.7) in the amoxicillin group (Mean Difference - 0.1, 95% CI -0.5 to 0.3, *P* = 0.63). Fewer adverse events were reported in the azithromycin group (Risk Ratio 0.50, 95% CI 0.31 to 0.82, *P* = 0.006).

**Conclusions:** No differences were found in weight gain between groups. Given the ability to administer a single dose and the potential for fewer adverse events, azithromycin may be an alternative to amoxicillin for uncomplicated SAM. With strong enrollment and follow-up, a larger trial in this setting is feasible.

## INTRODUCTION

Severe acute malnutrition (SAM) is estimated to impact 19 million children under 5 each year and is responsible for 7% of mortality in this age group.^1^ Infectious disease is common in children with SAM, who face an increased risk of mortality from diarrhea, measles, and pneumonia.^2 3^ However, identifying infection in this population can be challenging, since SAM suppresses the immune response, resulting in poor correlation between clinical signs and the presence of infection.^3-5^ Given this potential for asymptomatic infection, the World Health Organization (WHO) recommends that children with uncomplicated SAM be treated with a broad-spectrum oral antibiotic in addition to ready-to-use therapeutic food (RUTF).^6^

Many national nutritional guidelines recommend oral amoxicillin in the management of uncomplicated SAM, but evidence on the role of amoxicillin in this population is mixed. A trial in Malawi randomized children to amoxicillin, cefdinir, or placebo for 7 days and found that both antibiotics increased nutritional recovery and decreased mortality relative to placebo.^7^ Another trial in Niger compared a 7-day course of amoxicillin to placebo and did not find a difference in nutritional recovery or mortality between treatment groups.^8^ Children receiving amoxicillin in the Niger trial were less likely to be transferred to inpatient care and experienced a shorter time to recovery compared to placebo.^8^ Both trials found that children who received antibiotics experienced increased weight gain compared to children receiving placebo. Meta-analyses of the amoxicillin groups from these trials have produced confidence intervals consistent with no effect of amoxicillin on recovery (lower bound of 1.00) as well as up to a 6% increase in recovery with amoxicillin compared to placebo (upper bounds of 1.05 and 1.06).^9 10^ Another meta-analysis pooled both antibiotics examined in the two studies and suggested a 6% increase in recovery in the groups receiving any antibiotic compared to placebo.^11^ The difference in results in these two trials may indicate that antibiotics may not be required for uncomplicated SAM in some settings or that antibiotics other than amoxicillin should be considered.

Azithromycin may provide several advantages over amoxicillin in the management of uncomplicated SAM. A cluster-randomized trial in Niger demonstrated a substantial reduction in mortality with a single dose of azithromycin at 20 mg/kg in children without established infection.^12^ A subgroup analysis of this trial suggested that the administration of a single dose of azithromycin to severely malnourished children may minimize the gap in mortality between malnourished children and their well-nourished peers.^13^ In addition, azithromycin has a long half-life, which may reduce the burden of infectious organisms for several weeks.^14^ Indeed, the reported duration of population-level reductions in infectious disease burden ranges from 2 weeks to 6 months after a single azithromycin distribution.^15-17^ With the potential for longer term protection, azithromycin could be administered during routine SAM outpatient visits as a single dose, removing the need to rely on caregivers to administer multiple doses.^18^ Finally, amoxicillin is more commonly used than macrolide antibiotics for routine treatment in many settings experiencing high malnutrition.^19 20^ The potential for the selection for antimicrobial resistance and thus reduced efficacy may be greater with amoxicillin compared to azithromycin.^21^

In this pilot randomized trial, we aimed to compare the effect of azithromycin to amoxicillin on weight gain and nutritional recovery in the management of uncomplicated SAM in Burkina Faso. We hypothesized that children randomized to receive azithromycin will have increased weight gain and increased nutritional recovery compared to amoxicillin 8 weeks after admission to the nutritional program. In addition, we aimed to establish the feasibility of conducting a full-scale randomized controlled trial to evaluate different antibiotics for uncomplicated SAM in this setting.

## METHODS

### Design overview

This individual-randomized trial was designed to establish the feasibility of a larger trial and to provide preliminary evidence on the effect of azithromycin compared to amoxicillin on weight gain and nutritional recovery in the management of uncomplicated SAM in children 6-59 months old in Burkina Faso. Eligible children presenting to nutritional programs in the study area were randomized to a single dose of oral azithromycin or a 7-day course of amoxicillin upon admission. All children received standard outpatient treatment for uncomplicated SAM according to Burkina Faso guidelines. Participants were followed weekly at routine follow-up visits until nutritional recovery and all participants were asked to return for a final study visit at 8 weeks. The study protocol has been previously published.^22^

### Patient involvement

Patients were not involved in the design or conduct of this research.

### Study setting and participants

Participants were enrolled at 6 Centre de Santé et de Promotion Sociale (CSPS) with nutritional programs for SAM from a catchment area of 54 communities in Boromo District, Burkina Faso. CSPSs are government-run primary health facilities that provide basic preventive care and treatment. All children under 5 years of age receive care free of charge subsidized by the government. Each enrollment site screened children presenting to the CSPS for eligibility. Community health workers also screened children in the study catchment areas for SAM using mid-upper arm circumference (MUAC). Children 6-59 months old living in the study area were eligible for enrollment if they presented with: a weight-for-height Z-score (WHZ) < -3 or MUAC < 11.5 cm; no edema; no antibiotic use or SAM treatment in the prior 7 days; no clinical complications requiring antibiotic or inpatient treatment; no congenital abnormality or chronic illness; sufficient appetite; and written informed consent.

### Randomization and masking

The randomization sequence was generated by the UCSF data team in R (R Foundation for Statistical Programming, Vienna, Austria) and implemented through an electronic data collection system. After a baseline assessment, enrolled children were randomized 1:1 to azithromycin or amoxicillin without stratification or blocking. Participants and study personnel administering treatment were not masked to treatment assignment given logistical constraints and the lack of placebo. Outcome data collectors were masked to treatment assignment. Allocation concealment from outcome assessors was achieved by restricting access to the randomization assignment and ensuring that a separate masked team member collected outcome data.

### Interventions

Azithromycin was administered as a single directly observed weight-based dose (20 mg/kg) at the time of enrollment (Azithrin oral suspension 200 mg/5 ml, Strides Shasun Ltd, Bangalore, India). Amoxicillin was administered as a 7-day course at a dosage of 80 mg/kg of body weight per day divided into 2 daily doses (Amoxicillin syrup 250 mg/5ml, Reyoung Pharmaceutical, Shandong, China) according to national guidelines.^23^ CSPS study personnel administered and directly observed the first dose of amoxicillin at enrollment and caregivers administered remaining doses at home. Apart from antibiotics, all enrolled children received standard outpatient treatment for uncomplicated SAM according to the guidelines of the government of Burkina Faso,^23^ which include provision of RUTF (Plumpy’Nut, Nutriset, Malaunay, France), anti-malarials if positive for malaria by rapid diagnostic test, anti-parasitics, missing vaccinations, and vitamin A supplementation. Each RUTF sachet contains 500 kcal, 12.8 g of protein, 30.3 g of lipids, 45 g of carbohydrates, and micronutrients. During the course of the trial, Burkina Faso faced a national shortage of RUTF, which impacted some participants.

Caregivers were instructed to report adverse events experienced within 7 days of enrollment. Follow-up visits included a survey of adverse events, including fever, diarrhea, vomiting, abdominal pain, skin rash, and constipation. Serious adverse events were defined as death, hospitalization, or any other life-threatening situation and were reported to the Medical Monitor within 24 hours to determine whether the event was possibly related to the study drug.

### Participant timeline and data collection

At enrollment, the baseline assessment involved a questionnaire on socioeconomic status and feeding practices in addition to anthropometric assessments, including height (ShorrBoard Infant/Child Measuring Board, Weight and Measure, LLC, USA), weight (Seca 874dr scale, Seca, Germany), and MUAC (Shorr Child MUAC tape, Weight and Measure, LLC, USA).

Children were followed weekly (±2 days) at routine follow-up visits until nutritional recovery and again at 8 weeks (−5 days, +21 days) after enrollment. Anthropometry, vital status, adverse events, and clinical examination outcomes were recorded at each visit. All data were collected electronically using a mobile application (CommCare by Dimagi, Cambridge, MA, USA) and uploaded to a secure, cloud-based server.

### Outcomes

The primary feasibility outcomes included 1) enrollment potential defined as average number of participants enrolled per site per day, 2) refusals calculated as the percentage of eligible participants refusing to participate, and 3) loss to follow-up defined as the percentage of enrolled participants with incomplete follow-up.

The primary clinical outcome was weight gain defined as grams per kilogram per day (g/kg/day) over the 8-week study period. Secondary outcomes included nutritional recovery by 8 weeks (WHZ ≥ -2 or MUAC ≥ 125 mm on two consecutive visits, based on the criterion used for enrollment), nonresponse at 8 weeks, transfer to inpatient care, mortality, clinical signs of infection, height-for-age Z-score (HAZ), MUAC, weight-for-age Z-score (WAZ), and WHZ. Anthropometric outcomes were defined according to the 2006 World Health Organization Child Growth Standards.^24^ Sensitivity analyses included the use of alternative definitions for nutritional recovery. One sensitivity analysis defined nutritional recovery as WHZ ≥ -2 or MUAC ≥ 125 mm on two consecutive visits or WHZ ≥ -2 or MUAC ≥ 125 mm at the final study visit. Another sensitivity analysis compared time to recovery by group.

### Sample size and statistical considerations

As a pilot trial, a sample size of 300 (150 per group) was chosen pragmatically to balance resource constraints against the objectives of the trial. Given this sample size, we estimated having 80% power to detect a 27% increase in weight gain (g/kg/day) in children receiving azithromycin compared to amoxicillin. Assumptions for this sample size calculation were based on estimates from a trial comparing amoxicillin to placebo for children with uncomplicated SAM in Niger.^8^ These assumptions include an average weight gain of 4.9 g/kg/day over 8 weeks in the amoxicillin group, a standard deviation of 3.9 g/kg/day, loss to follow-up of 10%, and an alpha of 0.05.

Feasibility outcomes were summarized descriptively in aggregate and by group. The primary clinical outcome analysis compared weight gain by group from baseline to 8 weeks using a linear regression model with treatment group as the sole covariate. Similar sensitivity analyses compared period-specific weight gain by group at from week 0-1, 1-2, 2-4, and 4-8. Secondary binary outcomes like nutritional recovery and clinical outcomes were analyzed with log-binomial regression. Modified Poisson models with robust standard errors were used if the log-binomial models failed to converge.^25^ Other secondary anthropometric outcomes (weight, height, MUAC, WAZ, WHZ, HAZ) were analyzed as continuous variables at all time points and overall using linear regression models adjusted for baseline measurements. Time to event outcomes were compared by group using Cox proportional hazards regression. No adjustments were made for multiple comparisons as all outcomes were pre-specified. All analyses were intention-to-treat and conducted in Stata 15.1 (StataCorp, College Station, TX) or R (R Foundation for Statistical Programming, Vienna, Austria).

### Ethical approval and trial oversight

Ethical approval was obtained from the Comité d’Ethique du Burkina Faso and the University of California, San Francisco before study activities began. Written informed consent was obtained from a caregiver for the participation of each child. An infectious disease specialist at UCSF served as the Medical Monitor to provide clinical oversight on the study design and to monitor serious adverse events. A Data and Safety Monitoring Committee (DSMC) approved the protocol before the study began and monitored study progress, data collection, and adverse events through quarterly progress reports and annual meetings with the study team.

## RESULTS

Between 3 June 2020 and 9 October 2020, 312 children presenting to enrollment sites were screened for eligibility and 301 were enrolled (Figure 1), resulting in an average of > 2 participants enrolled overall per day or almost 1 participant enrolled per site per day. No caregivers refused to have their child participate. Of those enrolled, 161 were randomized to receive a single dose of oral azithromycin and 140 to a 7-day course of oral amoxicillin. Two hundred eighty-two children (93.7%) completed the 8-week follow-up visit and were included in the primary analysis, with 150 (93.2%) in the azithromycin group and 132 (94.2%) in the amoxicillin group. Overall, baseline characteristics were balanced by treatment group (Table 1). In both groups at baseline, 164 (54.5%) of children presented with a WHZ < -3 and 243 (80.7%) presented with MUAC < 11.5 cm.

**Table 1.** Baseline characteristics by treatment group.^**a**^. Demographic, household, and anthropometric characteristics of enrolled participants at the initial study visit.

| Characteristic | Total<br>(N=301) | Azithromycin<br>(N=161) | Amoxicillin<br>(N=140) |
| --- | --- | --- | --- |
| Enrollment site (n, %) |  |  |  |
| *Boromo* | 48 (16.0%) | 27 (16.8%) | 21 (15.0%) |
| *Farah* | 74 (24.6%) | 42 (26.1%) | 32 (22.9%) |
| *Ouahabou* | 41 (13.6%) | 19 (11.8%) | 22 (15.7%) |
| *Pa* | 54 (17.9%) | 26 (16.2%) | 28 (20.0%) |
| *Poura* | 34 (11.3%) | 18 (11.2%) | 16 (11.4%) |
| *Siby* | 50 (16.6%) | 29 (18.0%) | 21 (15.0%) |
| Enrollment criterion<sup>b</sup> |  |  |  |
| *MUAC < 11.5 cm only* | 101 (33.6%) | 62 (38.5%) | 39 (27.9%) |
| *WHZ < -3 SD only* | 40 (13.3%) | 17 (10.6%) | 23 (16.4%) |
| *Both MUAC < 11.5 cm and WHZ < 3 SD* | 160 (53.2%) | 82 (50.9%) | 78 (55.7%) |
| Age, months (mean, SD) | 16.7 (8.4) | 16.9 (8.4) | 16.5 (8.2) |
| Female sex (N, %) | 169 (56.2%) | 92 (57.1%) | 77 (55.0%) |
| Currently breastfeeding (N, %) | 212 (70.4%) | 108 (67.1%) | 104 (74.3%) |
| Exclusively breastfeeding (N, %) | 21 (7.0%) | 10 (6.2%) | 11 (7.9%) |
| No. of children under 5 in the household<br>(mean, SD) | 1.9 (1.6) | 2 (1.7) | 1.9 (1.5) |
| Household water source (N, %) |  |  |  |
| *Running water* | 97 (32.2%) | 50 (31.1%) | 47 (33.6%) |
| *Protected well* | 121 (40.2%) | 68 (42.2%) | 53 (37.9%) |
| *Unprotected well* | 29 (9.6%) | 18 (11.2%) | 11 (7.9%) |
| *Well (not specified)* | 54 (17.9%) | 26 (16.2%) | 28 (20.0%) |
| Household sanitation (N, %) |  |  |  |
| *Flush toilets* | 26 (8.6%) | 14 (8.7%) | 12 (8.6%) |
| *Improved pit latrine* | 10 (3.3%) | 4 (2.5%) | 6 (4.3%) |
| *Slab latrine* | 138 (45.9%) | 67 (41.6%) | 71 (50.7%) |
| *Unimproved latrine* | 81 (26.9%) | 52 (32.3%) | 29 (20.7%) |
| *Open defecation* | 46 (15.3%) | 24 (14.9%) | 22 (15.7%) |
| Mother's age, years (mean, SD) | 26.1 (6.1) | 26.0 (6.2) | 26.2 (6.0) |
| Literate mother (N, %) | 54 (17.9%) | 30 (18.6%) | 24 (17.1%) |
| No. times mother pregnant (mean, SD) | 2.3 (2.0) | 2.2 (1.9) | 2.5 (2.1) |
| No. of mother's children still alive (mean,<br>SD) | 1.9 (1.6) | 1.8 (1.5) | 2.0 (1.8) |
| Weight, kg (mean, SD) | 6.7 (1.3) | 6.8 (1.3) | 6.6 (1.3) |
| Height, cm (mean, SD) | 71.9 (6.6) | 72.2 (6.6) | 71.6 (6.6) |
| HAZ |  |  |  |
| *Mean score (mean, SD)* | -2.4 (1.6) | -2.4 (1.5) | -2.5 (1.6) |
| *HAZ < -3 (N, %)* | 113 (37.5%) | 55 (34.2%) | 58 (41.4%) |
| MUAC |  |  |  |
| *Mean circumference, cm (mean, SD)* | 11.2 (0.5) | 11.2 (0.4) | 11.2 (0.5) |
| *< 11.5 cm (N, %)* | 243 (80.7%) | 136 (84.5%) | 107 (76.4%) |
| WAZ |  |  |  |
| *Mean score (mean, SD)* | -3.5 (1.1) | -3.4 (1.0) | -3.5 (1.1) |
| *Score below -3 (N, %)* | 211 (70.1%) | 115 (71.4%) | 96 (68.6%) |
| WHZ |  |  |  |
| *Mean score (mean, SD)* | -3.1 (1.0) | -3.0 (1.1) | -3.1 (1.0) |
| *Score below -3 (N, %)* | 164 (54.5%) | 84 (52.2%) | 80 (57.1%) |

<sup>a</sup>Percentages may not total to 100% due to rounding
<sup>b</sup>MUAC and WHZ assessments to determine eligibility occurred before the baseline anthropometric assessments. Numbers of children meeting MUAC and/or WHZ thresholds may differ between these two timepoints (eligibility screening and baseline anthropometric assessment).

**Figure 1.**
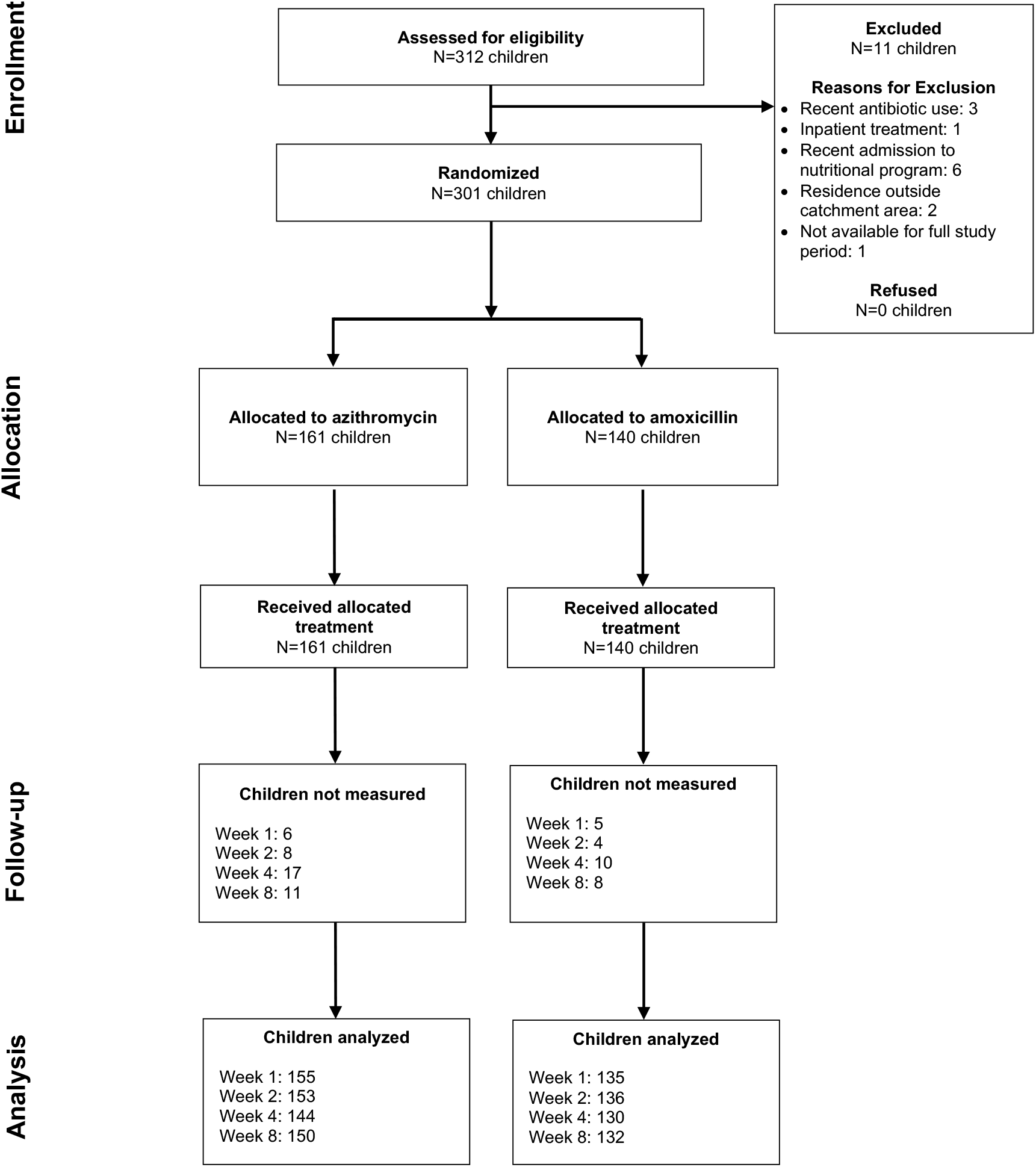
Flow of study participants. CONSORT diagram of flow of study participants from enrollment through analysis by treatment group.

Weight gain velocity (g/kg/day) is summarized by group overall (primary outcome) and by time point in Table 2 and Figure 2. On average, children in both treatment groups gained weight at all time points, with the greatest weight gain in the first week (Table 2). Average weight gain over the 8-week period was 2.5 g/kg/day (SD 2.0) in the azithromycin group and 2.6 g/kg/day (SD 1.7) in the amoxicillin group. At the final study visit, there was no difference in weight gain when comparing groups (Mean Difference -0.1, 95% CI -0.5 to 0.3, *P* = 0.63). No differences were demonstrated in weight gain at individual time points or in other anthropometric outcomes when compared by treatment group (Table 2).

**Table 2.**
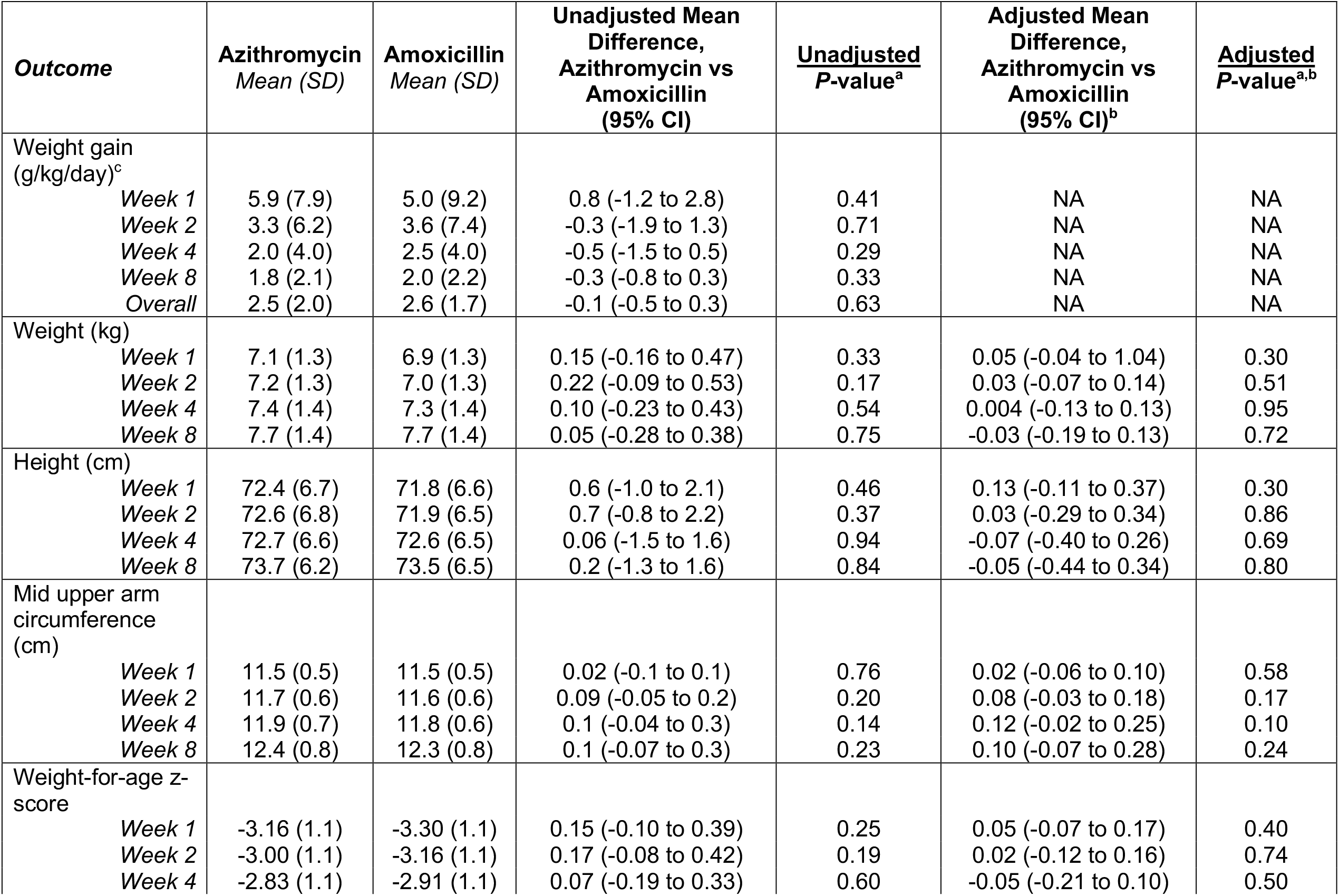
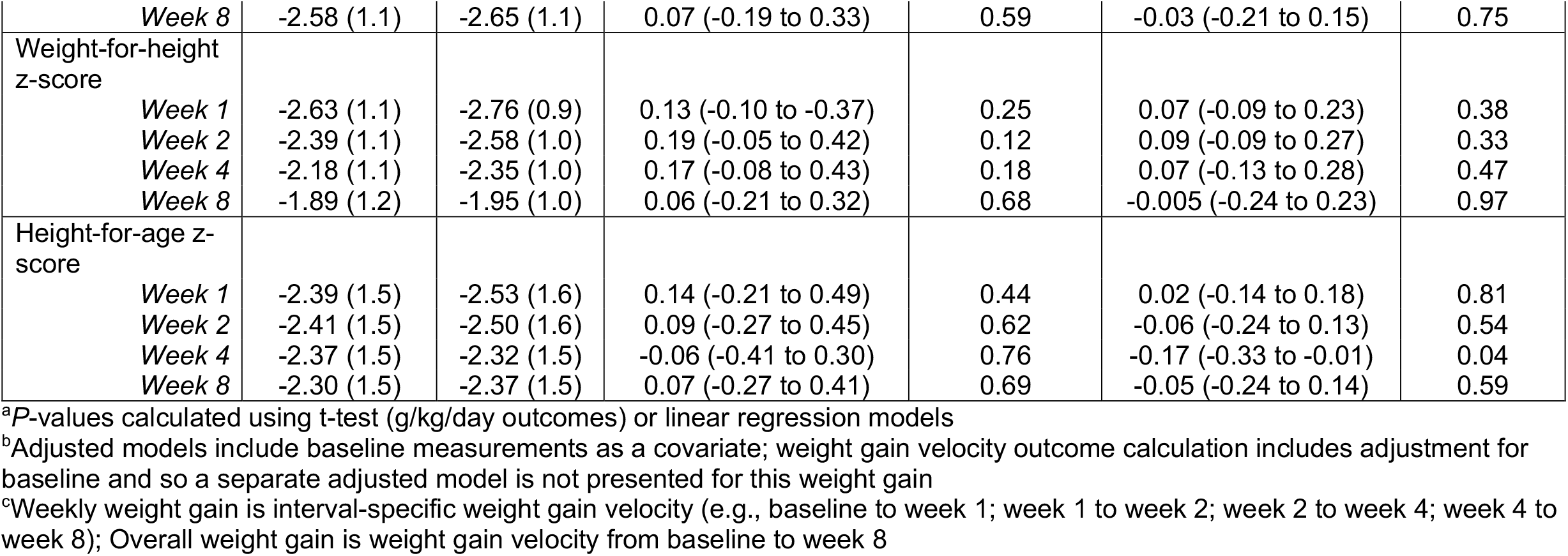
Longitudinal anthropometric outcomes by treatment group.

**Figure 2.**
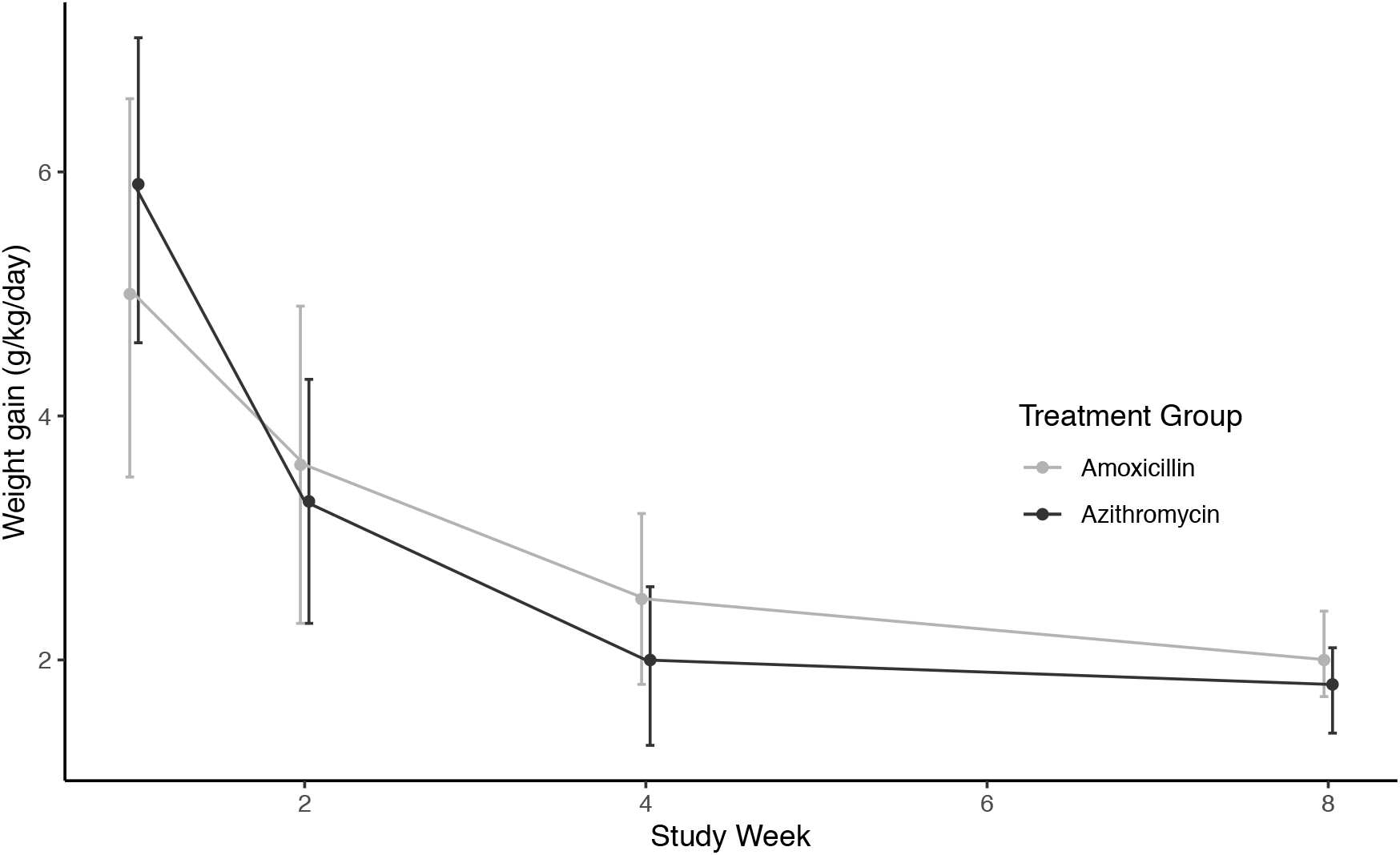
Weight gain velocity in g/kg/day by treatment interval and treatment group. Week 1 indicates weight gain (g/kg/day) from Baseline to Week 1, Week 2 is Week 1 to Week 2, and so on. All children had measurements collected at Baseline, Weeks 1-4, and Week 8. Measurements were only collected on Weeks 5-7 for children who had not recovered and thus not presented.

By 8 weeks, 56 children (37.3%) in the azithromycin group and 48 children (36.4%) in the amoxicillin group met the definition for nutritional recovery (Table 3; Risk Ratio 1.03, 95% CI 0.76 to 1.40, *P* = 0.87). In a sensitivity analysis that also included children who met the definition for recovery at the final visit, 98 children (65.3%) in the azithromycin group and 87 children (65.9%) in the amoxicillin group had recovered. The sensitivity analysis also found no difference in nutritional recovery by treatment group (Table 3; Risk Ratio 0.99, 95% CI 0.84 to 1.18, *P* = 0.92). Similarly, additional sensitivity analyses found no difference in time to nutritional recovery by group with either definition of nutritional recovery (original definition: Hazard Ratio 1.05, 95% CI 0.71 to 1.53, *P* = 0.82; sensitivity analysis definition: Hazard Ratio 1.02, 95% CI 0.77 to 1.35, *P* = 0.90).

**Table 3.** Clinical outcomes and adverse events by treatment group.

| **Outcome** | **Azithromycin** | **Amoxicillin** | **Risk Ratio<sup>a</sup>**<br>**(95% CI)** | **P-value** |
| --- | --- | --- | --- | --- |
| Nutritional recovery, original definition<sup>b</sup> | 56 (37.3%) | 48 (36.4%) | 1.03 (0.76 to 1.40) | 0.87 |
| Nutritional recovery, sensitivity definition<sup>c</sup> | 98 (65.3%) | 87 (65.9%) | 0.99 (0.84 to 1.18) | 0.92 |
| Nonresponse at 8 weeks<sup>d</sup> | 89 (58.9%) | 80 (58.8%) | 1.00 (0.74 to 1.36) | 0.99 |
| Death | 2 (1.3%) | 1 (0.8%) | 1.75 (0.16 to 19.3) | 0.65 |
| Sought medical care by 8 weeks | 27 (17.3%) | 29 (21.0%) | 0.82 (0.49 to 1.39) | 0.47 |
| Hospitalized by 8 weeks | 1 (0.6%) | 1 (0.7%) | 0.88 (0.06 to 14.1) | 0.93 |
| Recent symptoms of infection, 2 weeks after treatment |  |  |  |  |
| 3+ watery stools in the last 24 h | 2 (1.3%) | 2 (1.5%) | 0.89 (0.13 to 6.31) | 0.91 |
| 14 days of diarrhea | 0 | 0 | N/A | N/A |
| Blood/mucus in stool | 0 | 0 | N/A | N/A |
| Difficulty breathing | 0 | 3 (2.2%) | N/A | N/A |
| Tachypnea<sup>e</sup> | 6 (3.9%) | 7 (5.2%) | 0.76 (0.26 to 2.27) | 0.63 |
| Adverse events, 2 weeks after treatment |  |  |  |  |
| Any | 21 (13.7%) | 37 (27.2%) | 0.50 (0.30 to 0.86) | 0.01 |
| Fever | 16 (10.5%) | 23 (16.9%) | 0.62 (0.33 to 1.17) | 0.14 |
| Diarrhea | 5 (3.3%) | 12 (8.8%) | 0.37 (0.13 to 1.05) | 0.06 |
| Vomiting | 5 (3.3%) | 4 (2.9%) | 1.11 (0.30 to 4.14) | 0.88 |
| Abdominal pain | 0 | 3 (2.2%) | N/A | N/A |
| Skin rash | 1 (0.7%) | 1 (0.7%) | 0.89 (0.06 to 14.2) | 0.93 |
| Constipation | 0 | 1 (0.7%) | N/A | N/A |

<sup>a</sup>Estimated using modified Poisson model with robust standard errors<sup>25</sup>
<sup>b</sup>Defined as MUAC > 12.5 cm and/or WHZ > -2 on two consecutive visits, corresponding to the admission criterion (MUAC and/or WHZ)
<sup>c</sup>Defined as MUAC > 12.5 cm and/or WHZ > -2 on two consecutive visits and/or at their final measurement
<sup>d</sup>Defined as MUAC < 12.5 cm and/or WHZ < -2 at the 8-week visit, corresponding to the admission criterion (MUAC and/or WHZ)
<sup>e</sup>Defined as a respiratory rate > 50 breaths/minute for children aged 6-11 months and > 40 breaths/minute for children aged 12-59 months

Of the 301 enrolled children, caregivers of 58 (19.3%) reported adverse events within the first 2 weeks of treatment. Adverse events were significantly different by treatment group, with 21 children (13.7%) in the azithromycin group and 37 children (27.2%) in the amoxicillin group experiencing any adverse event at 2 weeks after treatment (Table 3; *P =* 0.006). No differences were identified in comparisons of individual adverse events by group (Table 3). Overall, 3 serious adverse events were reported during the study period, all of which were deaths determined to be unrelated to the study drug. No differences were found in other clinical outcomes by treatment group, including nonresponse, death, and hospitalization (Table 3).

## DISCUSSION

In this pilot trial, we found no differences in weight gain or nutritional recovery between the azithromycin and amoxicillin groups. On average, children in both groups experienced 2.6 g/kg/day of weight gain and 37% of children reached nutritional recovery by 8 weeks, both of which are lower than expected by programs or reported in other studies. Nutritional programs target 8 g/kg/day of weight gain and expect > 75% of children with SAM to recover during management.^26^ Although other studies have also found lower weight gain and nutritional recovery in children with uncomplicated SAM in sub-Saharan Africa compared to program targets, these other study populations reported greater weight gain than this pilot trial and greater recovery proportions overall.^7 8 27^ In a sensitivity analysis, the proportion of children achieving nutritional recovery in this trial was more similar to those seen in other West African settings.^8 27^ The sensitivity analysis included children who met the recovery definition at the final visit rather than requiring two consecutive visits, suggesting that programmatic decisions about discharge and recovery may differ from the official guidelines. At the same time, Burkina Faso bears a particularly large burden of severe malnutrition which could underpin slower or lower recovery in this population.^1 28 29^ In this particular trial, the shortage of RUTF, a common occurrence in nutritional programs in this setting, could also have contributed to low weight gain and recovery.

Fewer adverse events were reported in children receiving azithromycin compared to amoxicillin. The difference appears to be driven by fewer events of diarrhea and fever in children receiving azithromycin, although these individual comparisons by group were not statistically significant at an alpha of 0.05. Azithromycin is known for its safety profile, has been well tolerated by children in community-based programs for trachoma and studies of child survival, and is known for its efficacy against common childhood infections.^12 14 30^ This finding is also consistent with results from a cluster-randomized trial that compared community distributions of azithromycin to placebo in children in Niger.^12^ In this setting, a survey of adverse events in infants after community drug distributions found fewer cases of diarrhea in children in communities receiving azithromycin compared to placebo (RR 0.68, 95% CI 0.49 to 0.96, *P* = 0.03).^31^ This trial also found evidence of a reduction in the relative abundance of a range of gut bacteria in children in communities receiving azithromycin compared to placebo,^32^ including pathogenic *Campylobacter* which is a common cause diarrheal disease in children.^33-35^

The results of this pilot trial establish the feasibility of a fully powered efficacy trial to evaluate antibiotics for uncomplicated SAM in this setting. Given the burden of malnutrition and the community-based presence of the nutritional programs in this district in Burkina Faso, this pilot enrolled 301 children in 6 sites over 4 months, confirming the potential ease and speed of enrollment in a larger trial. Nearly 94% of children completed the final 8-week follow-up visit and greater than 90% of children completed each interim follow-up visit. The excellent follow-up in this pilot reduces concerns about underpowered or biased outcome comparisons associated with loss to follow-up in a larger trial. In addition, the strong follow-up sets the stage for longer term follow-up time points in a future trial. Evidence indicates that post-discharge relapse is common, but research and treatment programs have typically focused on short-term outcomes during active management.^36^

The strengths of the study include the randomized design and the low loss to follow-up. Although a placebo was not used to maintain masking of participants or personnel administering treatment, outcome assessors remained masked, limiting the threat of bias in data collection. Limitations include the shortage of RUTF during enrollment, which could have influenced nutritional outcomes and contributed to the lower overall weight gain and recovery seen in this trial. While this impacts the broader generalizability of results, we do not believe the RUTF shortage affected treatment groups differentially. As this pilot aimed to determine the feasibility of conducting a larger trial, the trial was underpowered to detect a modest difference in nutritional outcomes in two groups receiving antibiotics. Previous trials have used nutritional recovery as a primary outcome to align with program targets, yet this dichotomized outcome may not fully capture the continuous recovery process experienced by malnourished children. As a continuous outcome, weight gain velocity would provide greater power to detect a difference in groups and may better characterize the dynamic nature of the treatment process. The results of this study may be generalizable to children with uncomplicated SAM in government-run nutritional programs in settings with a similar profile and burden of malnutrition.

## CONCLUSIONS

Overall, the results of this pilot study establish the feasibility and rationale for conducting a randomized controlled trial to evaluate antibiotics for uncomplicated SAM in this setting. Although we were unable to demonstrate a difference in nutritional outcomes by group, both groups experienced weight gain and nutritional recovery and the azithromycin group experienced fewer adverse events. Given its ease of dosing and safety profile, azithromycin may be a viable alternative antibiotic to consider in the adjunctive treatment of uncomplicated SAM.

## Data Availability

Deidentified individual participant data (including data dictionaries) will be made available, in addition to study protocols, the statistical analysis plan, and the informed consent form. The data will be made available upon publication to researchers who provide a methodologically sound proposal for use in achieving the goals of the approved proposal. Proposals should be submitted to.

## Acknowledgements

The authors would like to thank the study teams at the CSPS enrollment sites for their contributions to this trial. The authors would also like to thank the Data and Safety Monitoring Committee: Nisha Acharya (chair; MD, University of California, San Francisco), Emily Smith (ScD, MPH, George Washington University), and Christine Stewart (PhD, University of California, Davis).

## Funding/Support

This work was supported by the National Institutes of Health – National Institute of Child Health and Development (R21HD100932) and a UCSF REAC award.

### Clinical Trial Registration

ClinicalTrials.gov (NCT03568643); https://clinicaltrials.gov/ct2/show/NCT03568643

---

**Supplementary Table 1.** CONSORT Checklist.

| Section/Topic | Item | Checklist item | Reported in Section |
| --- | --- | --- | --- |
| **Title and abstract** |  |  |  |
|  | 1a | Identification as a randomised trial in the title | Title |
|  | 1b | Structured summary of trial design, methods, results, and conclusions | Abstract |
| **Introduction** |  |  |  |
| Background and objectives | 2a | Scientific background and explanation of rationale | Introduction |
|  | 2b | Specific objectives or hypotheses | Introduction |
| **Methods** |  |  |  |
| Trial design | 3a | Description of trial design (such as parallel, factorial) including allocation ratio | Methods: Design overview, Randomization and masking |
|  | 3b | Important changes to methods after trial commencement (such as eligibility criteria), with reasons | NA |
| Participants | 4a | Eligibility criteria for participants | Methods: Study setting and participants |
|  | 4b | Settings and locations where the data were collected | Methods: Study setting and participants |
| Interventions | 5 | The interventions for each group with sufficient details to allow replication, including how and when they were actually administered | Methods: Interventions |
| Outcomes | 6a | Completely defined pre-specified primary and secondary outcome measures, including how and when they were assessed | Methods: Outcomes |
|  | 6b | Any changes to trial outcomes after the trial commenced, with reasons | NA |
| Sample size | 7a | How sample size was determined | Methods: Sample size and statistical considerations |
|  | 7b | When applicable, explanation of any interim analyses and stopping guidelines | NA |
| Randomisation: |  |  |  |
| Sequence generation | 8a | Method used to generate the random allocation sequence | Methods: Randomization and masking |
|  | 8b | Type of randomisation; details of any restriction (such as blocking and block size) | Methods:<br>Randomization and masking |
| Allocation concealment mechanism | 9 | Mechanism used to implement the random allocation sequence (such as sequentially numbered containers), describing any steps taken to conceal the sequence until interventions were assigned | Methods:<br>Randomization and masking |
| Implementation | 10 | Who generated the random allocation sequence, who enrolled participants, and who assigned participants to interventions | Methods:<br>Randomization and masking |
| Blinding | 11a | If done, who was blinded after assignment to interventions (for example, participants, care providers, those assessing outcomes) and how | Methods:<br>Randomization and masking |
|  | 11b | If relevant, description of the similarity of interventions | Methods:<br>Randomization and masking;<br>Interventions |
| Statistical methods | 12a | Statistical methods used to compare groups for primary and secondary outcomes | Methods:<br>Sample size and statistical considerations |
|  | 12b | Methods for additional analyses, such as subgroup analyses and adjusted analyses | Methods:<br>Sample size and statistical considerations |
| **Results** |  |  |  |
| Participant flow (a diagram is strongly recommended) | 13a | For each group, the numbers of participants who were randomly assigned, received intended treatment, and were analysed for the primary outcome | Results; Figure 1; Table 1 |
|  | 13b | For each group, losses and exclusions after randomisation, together with reasons | Results; Figure 1 |
| Recruitment | 14a | Dates defining the periods of recruitment and follow-up | Results |
|  | 14b | Why the trial ended or was stopped | NA |
| Baseline data | 15 | A table showing baseline demographic and clinical characteristics for each group | Table 1 |
| Numbers analysed | 16 | For each group, number of participants (denominator) included in each analysis and whether the analysis was by original assigned groups | Results |
| Outcomes and estimation | 17a | For each primary and secondary outcome, results for each group, and the estimated effect size and its precision (such as 95% confidence interval) | Results; Table 1; Table 2 |
|  | 17b | For binary outcomes, presentation of both absolute and relative effect sizes is recommended | NA |
| Ancillary analyses | 18 | Results of any other analyses performed, including subgroup analyses and adjusted analyses, distinguishing pre-specified from exploratory | Results; Table 2; Table 3 |
| Harms | 19 | All important harms or unintended effects in each group (for specific guidance see CONSORT for harms) |  |
| **Discussion** |  |  |  |
| Limitations | 20 | Trial limitations, addressing sources of potential bias, imprecision, and, if relevant, multiplicity of analyses | Discussion |
| Generalisability | 21 | Generalisability (external validity, applicability) of the trial findings | Discussion |
| Interpretation | 22 | Interpretation consistent with results, balancing benefits and harms, and considering other relevant evidence | Discussion |
| **Other information** |  |  |  |
| Registration | 23 | Registration number and name of trial registry | Trial registration |
| Protocol | 24 | Where the full trial protocol can be accessed, if available | Methods |
| Funding | 25 | Sources of funding and other support (such as supply of drugs), role of funders | Funding/support |

